# Plasma Proteomics Linking Primary and Secondary diseases: Insights into Molecular Mediation from UK Biobank Data

**DOI:** 10.1101/2025.08.29.25334726

**Authors:** Hanyu Qian, Chao Wu, Bo Li, Anthony Rosenzweig, Meng Wang

## Abstract

Diabetes, hypertension, and dyslipidemia are major risk factors for cardiovascular, neurological, renal, and pulmonary diseases, yet clinically accessible molecular mediators linking these cardiometabolic conditions to downstream complications remain unclear. We analyzed plasma proteomic data from 53,030 UK Biobank participants with longitudinal follow-up. Mediation analysis evaluated circulating proteins linking three primary cardiometabolic diseases to 18 secondary outcomes. Mendelian randomization assessed potential causal relationships, and machine learning evaluated the predictive value of identified mediators. We identified 998 significant mediation pathways involving 337 unique plasma proteins. GDF15 consistently mediated associations between diabetes and cardiovascular diseases, and ACE2 linked poorly controlled diabetes to increased risk of nerve root and plexus disorders. Mediators were enriched in receptor-mediated signaling and molecular interaction pathways. Mendelian randomization supported potential causal roles for 44 proteins. Incorporating mediator proteins into machine learning models improved prediction of secondary disease risk beyond traditional clinical factors and other plasma proteins. Plasma proteins help mediate progression from cardiometabolic diseases to downstream complications. These findings provide a molecular map of disease mediation pathways, nominate biomarkers and therapeutic targets, and support improved risk stratification and targeted intervention.

## Introduction

With global populations expanding and aging, alongside the rising epidemic of metabolic disorders, the burden of chronic diseases is escalating at an unprecedented rate. Diabetes, hypertension, and dyslipidemia are among the most prevalent and consequential chronic health conditions worldwide. According to the World Health Organization (WHO), an estimated 1.28 billion adults are affected by hypertension, 830 million are living with diabetes, and approximately 39% of the global adult population exhibit elevated blood cholesterol levels^1^. These cardiometabolic chronic conditions, hereafter referred to as primary diseases, not only exert substantial economic and social burdens but also serve as major drivers of downstream health complications^2,3^. For example, diabetes and hypertension are well-recognized drivers of multi-organ disease development, contributing to elevated risks of cardiovascular outcomes—including heart failure (HF) and ischemic heart disease (IHD)^4–7^ —as well as renal dysfunction and chronic kidney disease^8^.

Previous evidence has identified a range of biological mediators linking metabolic and hemodynamic disorders (i.e. diabetes mellitus, hypertension, and dyslipidemia), to downstream pathological processes across multiple organ systems. These mediators encompass chronic inflammation, oxidative stress, endothelial dysfunction, altered adipokine signaling, and broader metabolic dysregulation. Pro-inflammatory signals, such as interleukin-6 (IL-6) and monocyte chemoattractant protein-1 (MCP-1) are consistently upregulated in metabolic disease and contribute to endothelial activation, monocyte recruitment, and extracellular matrix remodeling^9^. Concurrently, renin-angiotensin-aldosterone system (RAAS) activation enhances oxidative stress via NADPH oxidase, activates NF-κB in vascular and immune cells, leading to upregulation of inflammatory cytokines, chemokines, and adhesion molecules. These pathways accelerate hypertension, vascular inflammation, and atherosclerosis, especially in the context of diabetic and dyslipidemia^10,11^. Understanding these mediators not only advances mechanistic insight into the metabolic disease continuum but also highlights novel therapeutic targets beyond traditional risk factor management. However, many mediators identified to date may be challenging to assess routinely in clinical practice due to limitations in measurement techniques or biomarker availability. Moreover, the broader landscape of clinically accessible mediators detectable in blood remains largely unexplored. A systematic characterization of such biomarkers could substantially improve risk stratification, refine preventive strategies, and facilitate the development of targeted interventions, particularly for populations facing already high burdens of disease.

Emerging high-throughput plasma proteomic platforms, including Olink^12^ and SomaScan^13^, enable the quantification of thousands of plasma proteins from minimally invasive blood samples, providing unique insights into systemic biology. Plasma proteomics has shown promise in estimating organ biological aging^14,15^, predicting incident disease and mortality^16–18^, and linking diverse phenotypic traits to underlying disease status^19,20^. Leveraging these advances, we applied large-scale plasma proteomic profiling within a rigorous mediation analysis framework^21^ to systematically investigate the mediating roles of circulating proteins during disease development.

In this study, we used data from 53,030 UK Biobank participants (aged 40–69 years at baseline) with plasma proteomic measurements and longitudinal follow-up of clinical outcomes^22,23^ to identify mediating proteins in disease pathways linking three prevalent conditions—diabetes, hypertension, and dyslipidemia—to 18 secondary disease outcomes across four major clinical domains (Figure 1). We further annotated their biological functions, assessed their potential causal effects on secondary diseases using Mendelian randomization, and evaluated their value in predicting secondary disease risk among individuals with primary diseases. We also stratified participants by levels of primary disease control and severity, defined using continuous clinical biomarkers, to explore additional disease pathways and protein mediators.

**Figure 1.**
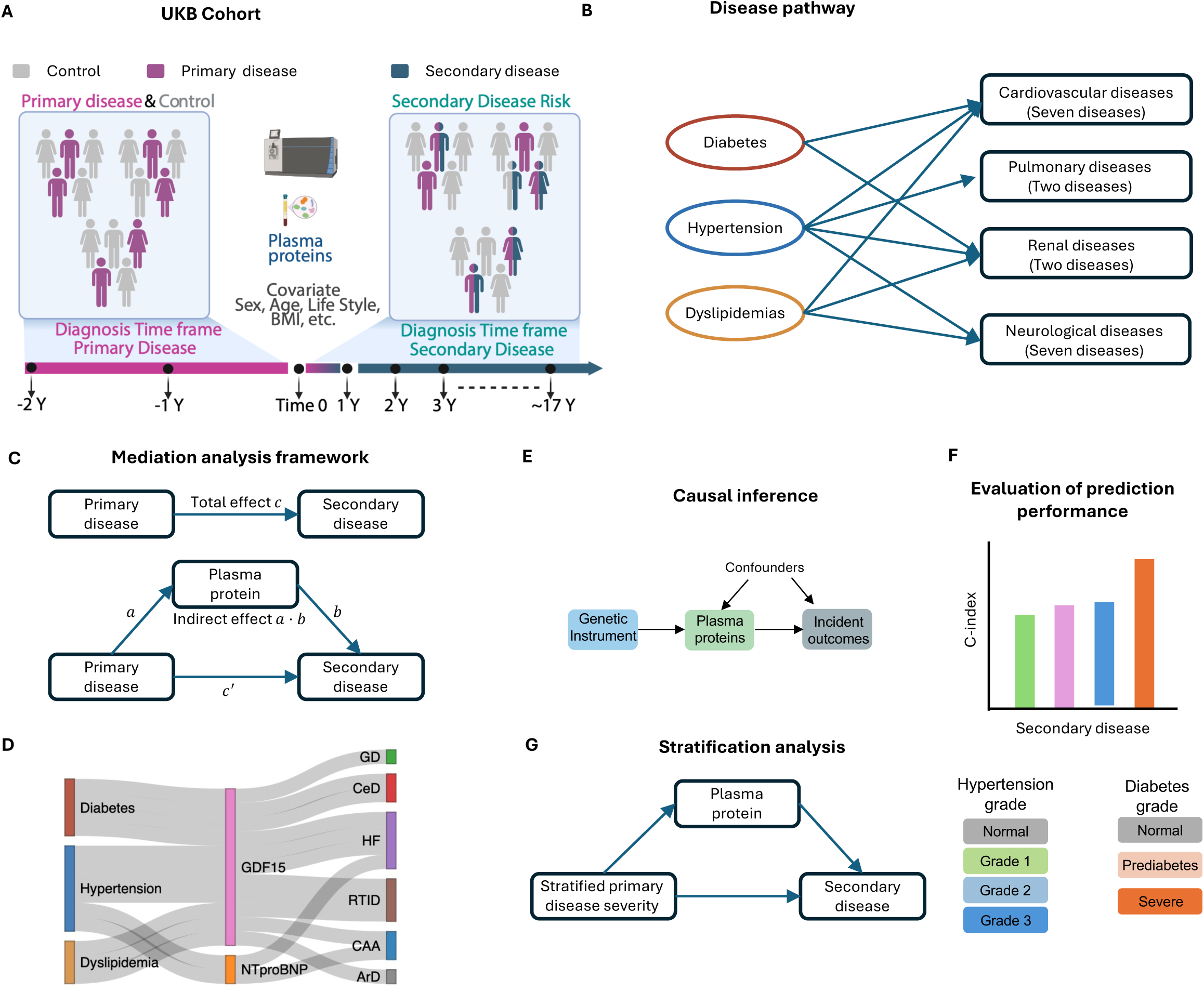
Overview of the study design and analytical framework. (A) Study cohort comprising 53,030 UK Biobank participants with plasma proteomic profiles and longitudinal clinical data. We used this cohort to investigate the mediating roles of plasma proteins linking primary diseases to secondary outcomes. (B) Disease pathways examined from three major cardiometabolic diseases—diabetes, hypertension, and dyslipidemia—to 18 secondary diseases across four clinical domains. (C) Mediation analysis framework, with primary disease status as the exposure, plasma protein expression as the mediator, and secondary disease risk as the outcome. (D) Examples of identified protein mediators linking primary diseases to secondary diseases. For instance, GDF15 and NT-proBNP act as key mediators bridging multiple disease pathways. (E) Causal inference framework using Mendelian randomization to evaluate the potential causal effects of identified protein mediators on secondary disease risk, with genetic variants as instrumental variables. (F) Illustration of the evaluation of prediction performance (C-index) using top-ranked protein mediators for secondary disease risk. (G) Stratification of primary diseases by severity and control (e.g., HbA1c and blood pressure), revealing additional protein-mediated disease pathways. (A) was created with BioRender.com.

## Results

### Population characteristics and diseases

A total of 53,030 participants were in the study, with an average age of 57.4 years, of whom 53.9% were women (Table S1). After data preprocessing (Methods), expression data for 2,922 plasma proteins and clinical data on three primary diseases--diabetes, hypertension, and dyslipidemia (Table S2), as well as 18 secondary diseases (Tables 1 and S3), were used for analysis to investigate candidate proteins involved in the primary-secondary disease pathway^23^. In this study, primary diseases were defined as chronic cardiometabolic conditions that increase the risk of subsequent health outcomes, while secondary diseases were defined as potential downstream outcomes associated with the presence of primary diseases. Primary disease status was determined based on whether participants were diagnosed with a primary disease within the period spanning two years before to one year after the plasma protein data collection. The number of positive cases per primary disease ranged from 1,053 (2.0%) to 3,012 (5.7%).

We examined 18 secondary diseases spanning four major organ systems: seven cardiovascular diseases (CVD), two renal diseases (RD), two pulmonary diseases (PuDs), and seven neurological diseases (NDs) (Tables 1 and S3). These domains were selected based on extensive evidence that the three primary cardiometabolic conditions—diabetes, hypertension, and dyslipidemia—exert systemic effects on the heart, kidney, lung, and brain^3,24–28^. Secondary disease outcomes were further selected according to the UK Biobank disease classification framework and required to have sufficient prevalence to ensure statistical power. Incident case counts, defined as diagnoses occurring after plasma protein collection, ranged from 106 (0.2%) to 8,962 (16.9%) across secondary diseases (Table S3). We randomly assigned 80% of participants to the training set to conduct mediation analyses and train prediction models to predict secondary disease risk based on the selected proteins, while the remaining 20% were used as the test set to evaluate out-of-sample predictive performance.

### Identification of diabetes-associated plasma protein mediating pathways

Diabetes was identified as a significant risk factor for four out of 18 secondary diseases, as indicated by hazard ratios (HRs) greater than 1 and Bonferroni adjusted *p* values29 < 0.05 in Cox regression models (Figures 2A and S1, Methods). The strongest associations were observed in glomerular diseases (GD; HR = 2.394, 95% confidence interval [CI]: 1.342 ∼ 4.271), and renal tubulo-interstitial disease (RTID; HR = 1.859, 95% CI: 1.555 ∼ 2.223), with both GD and RTID classified as renal diseases, consistent with prior reports^30–32^. Diabetes was also significantly associated with the risk of two CVDs, including cerebrovascular diseases (CeD) and HF, supporting previous studies^3^.

**Figure 2.**
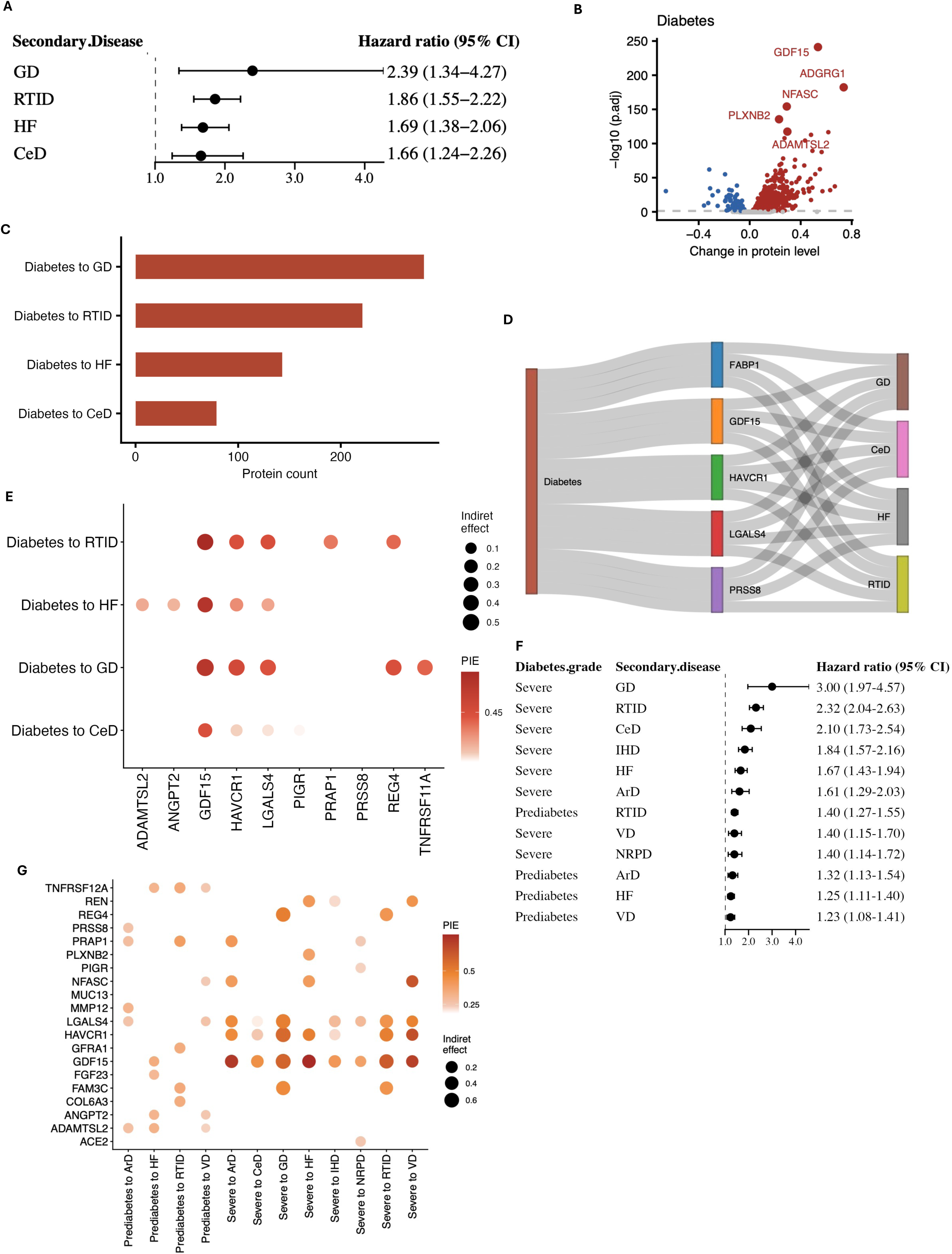
Results of diabetes-associated plasma protein mediating pathways. (A) Forest plot showing hazard ratios (HRs) with 95% confidence intervals (CIs) for secondary diseases associated with diagnosed diabetes, estimated using Cox proportional hazards models (only associations with *p* < 0.05 are shown). (B) Volcano plot showing changes in plasma protein expression associated with diabetes status and corresponding significance levels from linear models; the top five most significant proteins are highlighted. (C) Number of identified plasma protein mediators across diabetes-associated disease pathways. (D) Sankey diagram showing the top five protein mediators linking diabetes to the greatest number of secondary diseases. (E) Dot plot of the top five protein mediators ranked by indirect effect size in diabetes-associated pathways. Dot size represents the indirect effect, and dot color represents the proportional indirect effect (PIE). (F) Forest plot showing HRs with 95% CIs for secondary disease risk across three diabetes-control groups (only associations with *p* < 0.05 are shown). (G) Dot plot showing the top five protein mediators across diabetes-control groups identified through stratification analysis.

Among the 2,922 plasma proteins analyzed, 827 were significantly associated with diabetes (with Bonferroni adjusted *p* values < 0.05, Figure 2B). Several strong associations emerged including the association with GDF15, ADGRG1, and NFASC (*p* < 1 × 10 ^10^ for all). Notably, GDF15, a well-established metabolism-related protein, has been repeatedly linked to diabetes in prior studies^33^, supporting the robustness of our findings.

Significant diabetes-associated plasma proteins and secondary diseases were further examined under a mediation analysis framework^21^ to assess whether circulating proteins at least partially mediate the development from diabetes to its secondary outcomes (Figure 1C, Methods). To ensure robustness, we applied three criteria to identify mediators and their associated mediating disease pathways: (i) Bonferroni adjusted *p* < 0.05 for the indirect effect; (ii) An absolute indirect effect ≥ 0.01 (in terms of the log hazard ratio); and (iii) A proportional indirect effect (PIE), defined as the ratio of the indirect effect to the total effect, ≥ 10%. Overall, we identified 724 significant diabetes-associated protein mediation pathways involving 303 unique plasma proteins across four secondary diseases (Figure 2C and Table S4). Notably, 94.7% of these proteins were also independently associated with the corresponding secondary diseases after Bonferroni correction (Methods, Table S14). As expected, all indirect effects were positive, with HRs greater than 1, indicating that diabetes potentially increased the risk of secondary diseases via protein-mediated mechanisms.

Among the identified mediators, 68 proteins, including GDF15, LGALS4, PRSS8, and HAVCR1, mediated pathways linking diabetes to all four secondary diseases (Figures 2D and S2A). Many of these high-frequency mediators also exhibited strong and recurrent indirect effects across multiple disease pathways (Figure 2E and Table S5). GDF15, a well-established biomarker previously linked to diabetes^34,35^, emerged as the most prominent mediator, consistently showing the largest indirect effects across all four diabetes-related mediation pathways. In addition, LGALS4 ranked among the top five mediators by indirect effect size across diabetes-associated mediation pathways, consistent with previous observational studies reporting associations between circulating LGALS4 levels and diabetes-related traits^36^. The top-ranked mediator, defined as the protein with the largest indirect effect, remained unchanged in all four diabetes-related disease pathways in sensitivity analyses that incorporated ten additional covariates into the mediation framework, including the Townsend deprivation index (TDI), alanine aminotransferase, and other clinical factors. We then compared the top five mediating proteins identified in the sensitivity analyses with those from the primary analysis across all significant disease pathways. Overall, approximately 60% of the top-five proteins were shared between the two analyses, corresponding to an average Jaccard index^37^ of 0.443, indicating moderate-to-strong consistency in the prioritized mediator rankings despite additional covariate adjustment (Methods, Table S13).

Because glycemic control is a key determinant of diabetes-related complications, we further performed HbA1c-stratified analyses to complement the primary diagnosis-based analyses, examining whether glycated hemoglobin (HbA1c) levels were associated with differential risks of secondary diseases and distinct protein-mediated pathways. Participants were stratified according to HbA1c levels^38^: normal (HbA1c < 5.7%), prediabetes (5.7% ≤ HbA1c < 6.4%), and severe diabetes (HbA1c ≥ 6.4%). This stratified analysis identified four additional disease pathways that were not detected using diagnosis history alone (Figure 2F), including pathways linking diabetes to three cardiovascular diseases—IHD, arterial disease (ArD), and valve disorders (VD)—as well as one neurological disorder, nerve root and plexus disorders (NRPD). These findings suggest that poor glycemic control, even during the prediabetes stage, may increase the risk of downstream diseases.

We observed a substantial number of mediating proteins associated with prediabetes (Tables S10 and S11). Specifically, 345 proteins mediated the relationship between severe diabetes and eight secondary diseases, exceeding the number identified for prediabetes (232 proteins linked to 4 secondary diseases) and for diabetes defined by diagnosis history (303 proteins), with 162 proteins shared across these classifications (Figure S4). When comparing the magnitude of indirect effects for overlapping proteins linked to the same secondary diseases, severe diabetes consistently exhibited stronger mediation effects, followed by diabetes diagnosis history (Figure S4D).

Although most key protein mediators identified using HbA1c stratification overlapped with those identified using diagnosed history for the same secondary disease (Figure S4C, Tables S10 and S11), notable differences were observed (Figure 2G). For example, GDF15 and LGALS4 exhibited significant indirect effects across 12 disease pathways, representing the highest number of mediated pathways among all identified plasma proteins. Both proteins demonstrated strong indirect effects predominantly in the severe diabetes and diagnosed diabetes groups, but not in the prediabetes group (Figure 2H). In addition, stratification by HbA1c control level revealed additional pathway-specific mediators. For instance, ACE2 emerged as a strong mediator in the pathway leading to NRPD. Notably, ACE2 is a multifunctional metalloprotease with established roles in neuroprotection, neuronal development, and inflammatory regulation^39^.

### Identification of hypertension-associated plasma protein mediating pathways

We applied the mediation analysis framework to investigate the hypertension-related secondary diseases and their potential protein mediators (Methods). Hypertension was identified as a significant risk factor for five out of 18 secondary diseases in this study (Bonferroni adjusted *p* values < 0.05, Figure 3A). The strongest association was observed between hypertension and RTID (HR = 1.602, 95% CI: 1.374 ∼ 1.868). Hypertension was significantly associated with 548 plasma proteins (Bonferroni adjusted *p* values < 0.05, Figure 3B), with the strongest association observed for HAVCR1 (*p* < 1 × 10 ^10^).

**Figure 3.**
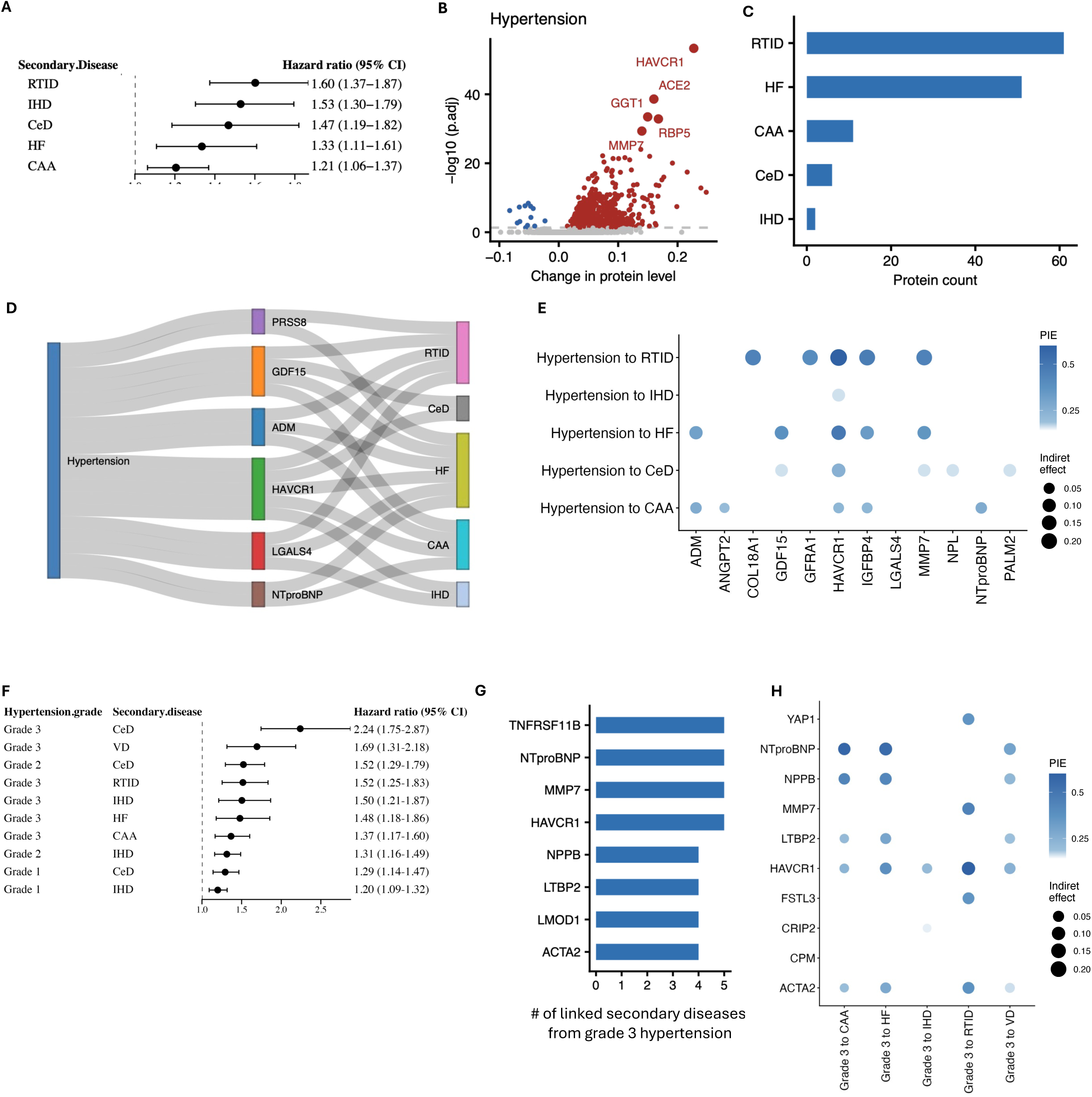
Results of hypertension-associated plasma protein mediating pathways. (A) Forest plot showing hazard ratios (HRs) with 95% confidence intervals (CIs) for secondary diseases associated with diagnosed hypertension, estimated using Cox proportional hazards models (only associations with *p* < 0.05 are shown). (B) Volcano plot showing changes in plasma protein expression associated with hypertension status and corresponding significance levels from linear models; the top five most significant proteins are highlighted. (C) Number of identified plasma protein mediators across hypertension-associated disease pathways. (D) Sankey diagram showing the top five protein mediators linking hypertension to the greatest number of secondary diseases. (E) Dot plot of the top five protein mediators ranked by indirect effect size in hypertension-associated pathways. Dot size represents the indirect effect, and dot color represents the proportional indirect effect (PIE). (F) Forest plot showing HRs with 95% CIs for secondary disease risk across three hypertension severity groups (only associations with p < 0.05 are shown). (G) Top protein mediators most frequently linking grade 3 hypertension to secondary diseases. (H) Dot plot of the top five protein mediators ranked by indirect effect in pathways linking grade 3 hypertension to secondary diseases.

Among these 548 hypertension-associated plasma proteins, 80 unique proteins exhibited significant indirect effects in at least one disease pathway (Figure 3C), of which 96.7% were independently significantly associated with secondary diseases. GDF15 and HAVCR1 emerged as key mediators, mediating four and five secondary disease associations, respectively (Figures 3D and S2B). The HAVCR1 gene encodes the immunoglobulin-mucin protein TIM-1/KIM-1, which is shed from injured proximal tubular cells and is currently recognized as an FDA-qualified, ultra-early biomarker for predicting both acute kidney injury and long-term decline in estimated glomerular filtration rate (eGFR)^40^. Another notable example was ADM which ranked among top five proteins mediating the associations between hypertension and two out of five secondary diseases (Figure 3E). ADM is markedly upregulated as a compensatory vasodilatory and natriuretic peptide that counterbalances excessive vascular tone, attenuates fibrosis, and preserves renal and cardiac function^41^. Elevated ADM levels have also been strongly associated with adverse cardiovascular and renal outcomes^42^, suggesting that the observed indirect effects may reflect sustained blood pressure elevation and compensatory vascular responses rather than direct promotion of disease development. Sensitivity analyses for hypertension showed consistent prioritization of top-ranked mediator proteins in all five pathways relative to the primary analysis, with moderate overlap in the top-five mediator lists across identified disease pathways, as indicated by an average Jaccard index of 0.43.

We further stratified participants according to systolic and diastolic blood pressure (SBP and DBP) into four categories: normal (< 140/90 mmHg), hypertension grade 1 (140–159/90–99 mmHg), hypertension grade 2 (160–179/100–109 mmHg), and hypertension grade 3 (≥ 180/110 mmHg), according to the 2023 European Society of Hypertension (ESH) guidelines^43^ (Methods). Using this stratification, we identified six significant disease pathways. In each pathway, participants in at least one hypertension grade, consistently including grade 3, exhibited a significantly higher risks of developing secondary diseases compared to those with normal blood pressure (Figure 3F). Five of these six secondary diseases overlapped with those identified using hypertension diagnosis history. The only additional disease development pathway was valve disorder (VD) among participants with grade 3 hypertension (HR = 1.694, 95% CI: 1.315–2.183), which was not observed among participants with grades 1 or 2 hypertension nor in analyses using diagnosis history. As expected, secondary diseases associated with milder hypertension were consistently retained among pathways identified at higher hypertension grades, with risk magnitude generally increasing with hypertension severity. Overall, the risk profile of participants with diagnosed hypertension generally resembled those with grade 2-3 hypertension.

From the mediation analysis, in total, 84 plasma proteins exhibited significant indirect effects across pathways from at least one hypertension grade group, with grade 3 hypertension displaying the broadest and strongest mediation signals (Figure S5). Participants with grade 3 hypertension were more likely to exhibit protein-mediated increases in secondary disease risk compared to those with lower hypertension levels, as evidenced by both a broader range of mediating proteins (78 unique proteins, Figure S5A) and stronger indirect effects through overlapping plasma proteins linked to the same secondary diseases (Figure S5B). Many of the key mediators linked to grade 3 hypertension overlapped with those identified when hypertension status was defined based on diagnostic history (Table S9), including HAVCR1, MMP7, and NT-proBNP (Figures 3G and 3H), in which NT-proBNP is a well-established biomarker of heart failure^44^.

Nevertheless, we also identified proteins uniquely associated with grade 3 hypertension that were not detected when using diagnostic history alone. For example, CRIP2, a protein highly expressed during cardiovascular development, emerged as a significant mediator in the grade 3 hypertension analysis but not under the diagnosis-based definition. Furthermore, comparative analyses of overlapping proteins linked to the same secondary diseases revealed that proteins identified through grade 3 hypertension consistently exhibited stronger indirect effects than those identified via hypertension diagnosis (Figure 5C).

### Identification of dyslipidemia-associated plasma protein mediating pathways

Dyslipidemia was identified as significant risk factor for three out of 18 secondary diseases (Bonferroni adjusted *p* values < 0.05, Figure 4A). In total, 258 circulating plasma proteins were significantly associated with dyslipidemia (Figure 4B). Among these, PCSK9 showed the strongest association (*p* < 1 × 10 ^10^), consistent with its well-established role in lipid metabolism and clinical intervention^45^. PCSK9 inhibitors, including monoclonal antibodies (alirocumab, evolocumab) and siRNA-based therapy (inclisiran), are approved adjuncts to statins and have demonstrated substantial efficacy in lowering LDL cholesterol levels and cardiovascular event rates in patients with familial hypercholesterolemia or established atherosclerotic cardiovascular disease (ASCVD)^45^.

**Figure 4.**
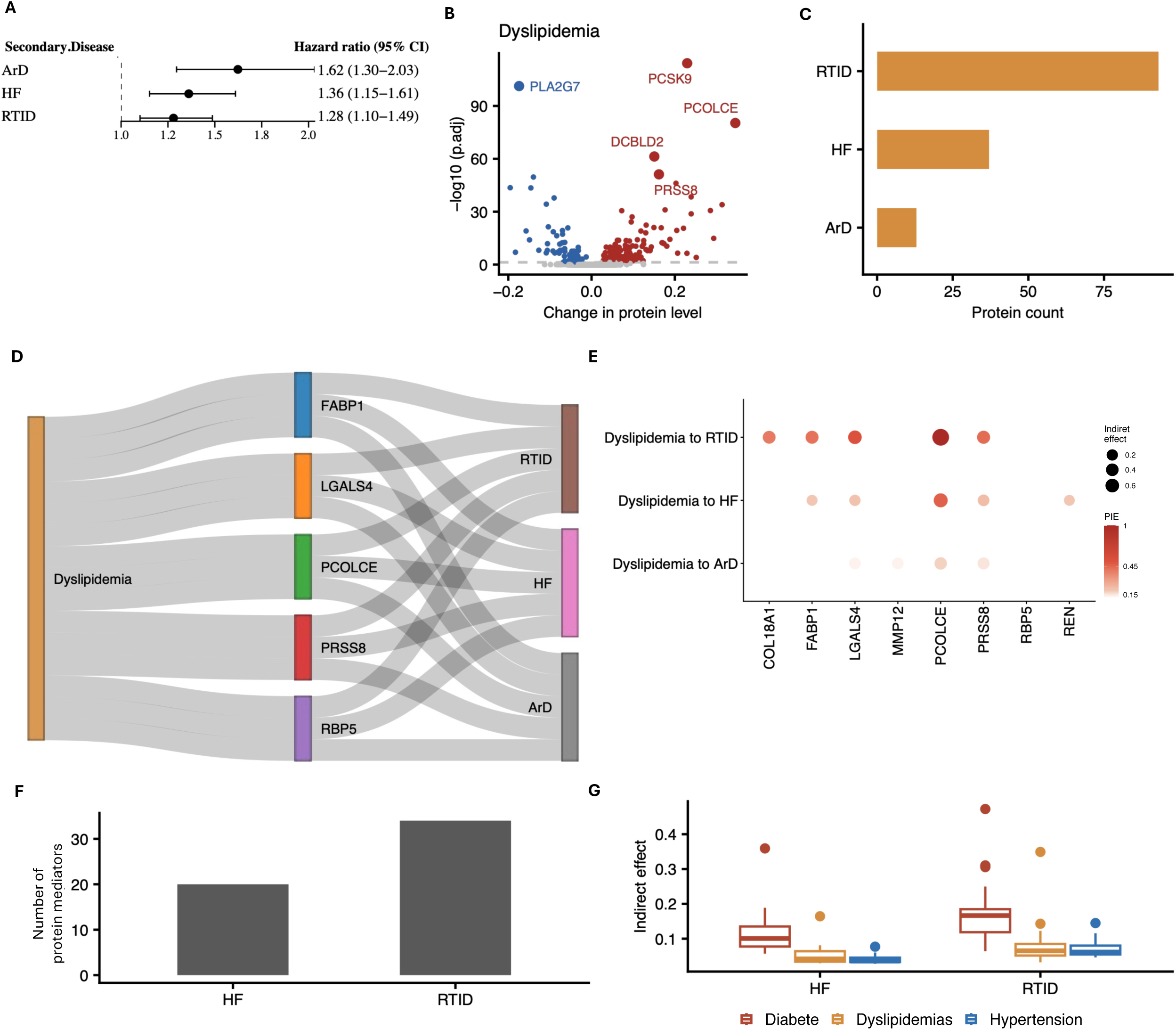
Results of dyslipidemia-associated plasma protein mediating pathways. (A) Forest plot showing hazard ratios (HRs) with 95% confidence intervals (CIs) for secondary diseases associated with diagnosed dyslipidemia, estimated using Cox proportional hazards models (only associations with *p* < 0.05 are shown). (B) Volcano plot showing changes in plasma protein expression associated with dyslipidemia status and corresponding significance levels from linear models; the top five most significant proteins are highlighted. (C) Number of identified plasma protein mediators across dyslipidemia-associated disease pathways. (D) Sankey diagram showing the top five protein mediators linking dyslipidemia to the greatest number of secondary diseases. (E) Dot plot of the top five protein mediators ranked by indirect effect size in dyslipidemia-associated pathways. Dot size represents the indirect effect, and dot color represents the proportional indirect effect (PIE). (F) Number of shared protein mediators linking all three primary diseases to HF and RTID. (G) Boxplot showing the distribution of indirect effects for these shared mediators across the three primary diseases.

Under the same mediation analysis framework, we identified 96 unique plasma proteins with significant indirect effects linking dyslipidemia to three secondary diseases (Figure 4C), of which 95.3% were also independently associated with the corresponding secondary diseases. Notably, 13 plasma proteins, including FABP1, LGALS4, exhibited significant indirect effects in all three pathways, respectively, representing the highest numbers among all dyslipidemia-associated mediators (Figures 4D and S2C). LGALS4 consistently ranked among the top five mediators by indirect effect across multiple dyslipidemia-associated pathways, suggesting a broader functional role in metabolic disease development. In addition to these shared mediators, FABP1, a key protein involved in fatty acid metabolism^46^, emerged as a prominent dyslipidemia-specific mediator (Figure 4E), underscoring its potential relevance to lipid-driven disease pathway. FABP1 acts at the interface of hepatic lipid handling and systemic metabolic homeostasis^29^. By regulating fatty acid trafficking and cholesterol metabolism, FABP1 influences the development of dyslipidemia, which in turn accelerates atherosclerosis, vascular dysfunction, and cardiac remodeling^47^. Elevated circulating FABP1 has been consistently linked to obesity, insulin resistance, and cardiovascular events^48^, raising the possibility that it operates as both a mechanistic mediator of lipid-driven pathology and a sensitive biomarker of hepatic stress. Sensitivity analyses for dyslipidemia also showed generally consistent mediator prioritization, with the top-ranked mediator remaining unchanged in two of the three identified disease pathways and moderate overlap in the top-five mediator lists compared with the primary analysis, as reflected by an average Jaccard index of 0.37.

We further identified 38 plasma proteins that exhibited significant indirect effects in pathways linking all three primary diseases to RTID or HF, three secondary diseases whose risks were significantly influenced by all primary conditions (Figures 4F, 4G and S3). Notably, GDF15 and IGFBP4 mediated associations across all three primary diseases and each of these secondary diseases, highlighting their central role in shared multimorbidity-related pathways. In addition, ACE2, a well-established therapeutic target in renal disease^49^, specifically mediated associations between all three primary diseases and RTID.

### Biological function annotation of the protein mediators

To characterize the biological functions underlying protein-mediated cross-disease relationship, we performed enrichment analysis using the Gene Ontology Biological Process (GO:BP)^50,51^ and KEGG^52^ databases to annotate enriched biological processes and pathways^53^ (Figure 5A, Table S12, and Methods).

**Figure 5.**
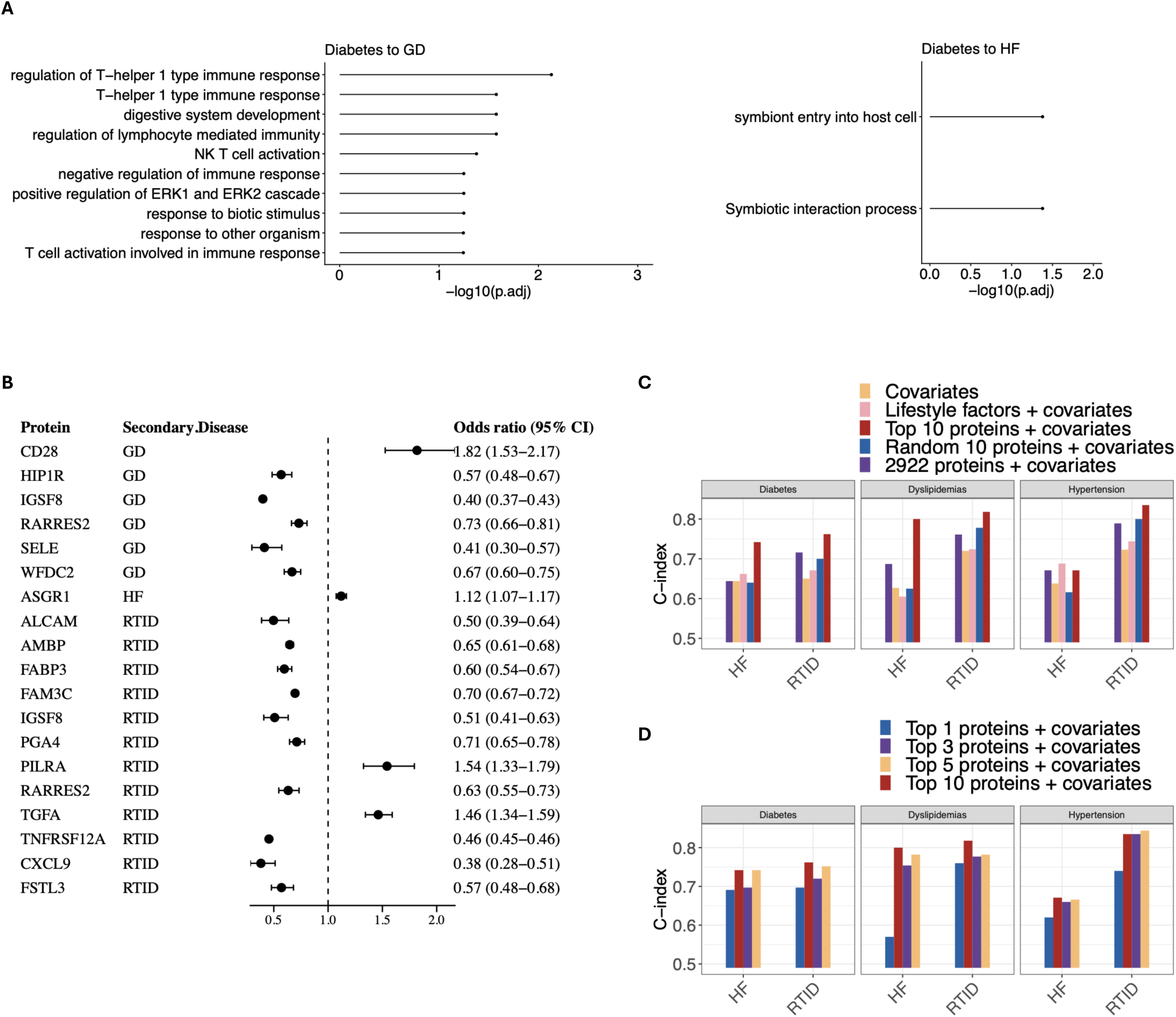
Biological function annotation of plasma protein mediators and evaluation of their causal and predictive effects. (A) Gene Ontology (GO) enrichment analysis of identified protein mediators across two representative pathways (full results are provided in the Table S12). (B) Forest plot showing odds ratios (ORs) with 95% CIs for secondary disease risk associated with protein mediators, as estimated by Mendelian randomization. (C) Comparison of prediction performance (C-index) for secondary disease risk across three primary diseases using different models: (i) all plasma proteins with covariates (age, sex, and BMI), (ii) covariates only, (iii) lifestyle factors and covariates, (iv) randomly selected sets of 10 plasma proteins with covariates (average performance over 100 replicates), and (v) the top 10 mediation-ranked protein mediators with covariates. (D) Comparison of prediction performance (C-index) using the top 1, 3, 5, and 10 mediation-ranked protein mediators with covariates.

**Table. 1.** Secondary Disease Names and Corresponding Abbreviations.

| Organ system | Disease name | Abbreviations |
| --- | --- | --- |
| Pulmonary diseases | Chronic lower respiratory diseases | CLRD |
| Pulmonary diseases | Other respiratory diseases principally affecting the interstitium | OPDRI |
| Renal diseases | Renal tubulo-interstitial diseases | RTID |
| Renal diseases | Glomerular diseases | GD |
| Neurological diseases | Systemic atrophies primarily affecting the central nervous system | SACNS |
| Neurological diseases | Parkison's disease | PD |
| Neurological diseases | Alzheimer's disease | AD |
| Neurological diseases | Multiple sclerosis | MS |
| Neurological diseases | Nerve root and plexus disorders | NRPD |
| Neurological diseases | Diseases of myoneural junction and muscle | MNJM |
| Neurological diseases | Other disorders of the nervous system | ODNS |
| Cardiovascular diseases | Ischaemic heart diseases | IHD |
| Cardiovascular diseases | Valve disorder | VD |
| Cardiovascular diseases | Myocardial Disorders | MyD |
| Cardiovascular diseases | Cardiac arrest and arrhythmias | CAA |
| Cardiovascular diseases | Heart failure | HF |
| Cardiovascular diseases | Cerebrovascular diseases | CeD |
| Cardiovascular diseases | Artery disease | ArD |

This enrichment was most pronounced in diabetes-to-GD mediation, which were significantly enriched for cytokine–cytokine receptor interaction. The corresponding GO:BP analysis identified regulation of T-helper-1 type immune response, regulation of lymphocyte-mediated immunity, and NK T-cell activation as leading processes, collectively implicating coordinated innate and adaptive immune activation in diabetic glomerular injury. Mediators linking diabetes with RTID were similarly enriched for cytokine–cytokine receptor interaction. By contrast, diabetes-to-HF mediation defined a distinct functional module centered on cell-entry and host–pathogen interactions (“symbiont entry into host cell” and “biological process involved in symbiotic interaction), driven by cell-surface entry and adhesion proteins including FURIN, ACE2, NECTIN2, ITGA5, EPHA2, ICAM1, and HAVCR1. This signature reflects enrichment of membrane receptor and adhesion machinery rather than the cytokine-driven immune programs characterizing the renal comparisons.

Collectively, these results indicate that immune signaling, particularly cytokine–cytokine receptor interaction and T-helper-1/lymphocyte-mediated immunity, constitutes the predominant biological process underlying protein-mediated diabetes-to-renal disease relationships, whereas diabetes-to-heart-failure mediation is distinguished by a discrete program of cell-surface receptor and adhesion proteins.

### Mendelian randomization reveals potential causal relationships between protein mediators and secondary diseases

We applied a Mendelian randomization^54^ (MR) framework^55,56^ to investigate whether plasma proteins exhibiting significant indirect effects also play causal roles in the development of secondary diseases. In this framework, protein abundance was treated as the exposure, and secondary disease incidence as the outcome, with genome-wide significant *cis*-protein quantitative trait loci (*cis*-pQTLs) as instrumental variables^23^. We employed a two-sample MR framework utilizing 12 methods to account for pleiotropy and heterogeneity, supplemented by Steiger directionality testing to confirm exposure-to-outcome effects^57^ (Methods).

Among the 303 protein mediators identified in pathways linking diabetes to four secondary diseases, 38 showed evidence of potential causal effects on four outcomes: CeD, GD, HF, and RTID. Similarly, 13 of 80 and 14 of 96 proteins associated with hypertension and dyslipidemia, respectively, had significant MR evidence of causal roles (Table S6). Among outcomes, RTID was associated with the largest number of putative causal proteins (25, Bonferroni adjusted^29^ *p* < 0.05).

Several findings recapitulated known disease mechanisms (Figure 5B), supporting the validity of the approach. ASGR1, a hepatic receptor central to LDL-cholesterol clearance and an established cardiovascular target, was inferred to causally increase risk of both heart failure (OR = 1.121*, p =* 1.01 × 10□□) and chronic renal failure (OR = 1.164, p *=* 2.79 × 10□^1^□), consistent with its dual role in lipid handling and cardiorenal injury^58,59^. The renin–angiotensin enzyme REN showed a strong causal association with chronic tubulo-interstitial nephritis (OR = 3.290, *p* = 3.95 × 10 □^23^), and the known kidney-function markers FGF23^60^ and UMOD^61^ (OR = 1.229*, p* = 1.71 × 10□^2^□) were likewise implicated in renal outcomes—reinforcing the biological plausibility of the mediator set.

The analysis also nominated proteins not classically linked to their outcomes. STC1 showed a pronounced causal association with chronic tubulo-interstitial nephritis (OR = 2.823, *p* < 1 × 10□^3^□□), and the tight-junction protein OCLN with chronic renal failure (OR = 4.771, *p* = 7.22 × 10□□^1^), suggesting calcium/phosphate signaling and epithelial barrier integrity as causal determinants rather than downstream markers. In the cerebrovascular and cardiac domains, UPB1 (cerebral infarction, OR = 0.993, *p* = 1.33 × 10□^2^□□) and LGALS4 (angina pectoris, OR = 1.005, *p* = 4.06 × 10 ²) emerged as candidates without prior description. Finally, several proteins acted pleiotropicall, EPHA2, IGSF8, RARRES2, WFDC2, and TGFA each reached significance for more than one outcome, indicating shared molecular nodes connecting the primary disease to multiple complications.

### Plasma protein mediators improve prediction of secondary disease risk among individuals with primary diseases

Using a LASSO-penalized Cox regression model^62^, we evaluated the predictive value of identified protein mediators for secondary disease risk among participants with a diagnosed primary disease. Analyses primarily focused on RTID and HF, which were associated with all three primary diseases, contained at least 10 proteins with significant mediation effects in the disease pathways, and had a sufficient number of positive cases to enable robust evaluation of predictive performance. For each disease pathway, participants with the corresponding primary disease were then selected from the previous defined training and testing sets, respectively. We used the 10-fold cross-validation to optimize hyperparameters within the corresponding training subset ^63^. Predictive performance was assessed using the concordance index^64^ (C-index) evaluated on held-out test subset. (Methods).

Across disease pathways, models incorporating the top ten mediation-ranked proteins consistently outperformed predictive models based on: (i) basic demographic covariates (age, sex, and body mass index), (ii) established lifestyle and clinical biomarkers (including blood pressure, cholesterol levels, smoking, and alcohol consumption), (iii) randomly selected sets of ten proteins (100 repeated samplings), and (iv) the full proteomic panel (Figure 5C and Table S7, Methods). All predictive models included basic covariates, including age, sex, and BMI. Notably, models incorporating either the entire proteomic profile or random subsets of proteins frequently showed lower predictive accuracy across multiple secondary diseases, indicating that inclusion of non-informative or weakly relevant proteins can dilute predictive signals, underscoring the importance of biologically informed feature selection for improving model performance.

We further investigated how the number of proteins included in the models affected prediction accuracy (Figure 5D). In five out of the six tested disease pathways, the model using the top 10 plasma proteins outperformed models using only the top five, three, or top one protein. However, in most cases, the predictive performance of models using the top five proteins achieved comparable performance, suggesting that a parsimonious set of high-impact proteins may yield clinically meaningful prediction while improving model interpretability and efficiency.

## Discussion

In this study, we systematically investigated the role of plasma proteins in mediating the development from three major cardiometabolic conditions—diabetes, hypertension, and dyslipidemia—to a broad range of downstream diseases using data from 53,030 UK Biobank participants. We identified 998 significant mediation pathways involving 337 unique plasma proteins across 18 secondary conditions, revealing both shared and disease-specific molecular mediators. Mendelian randomization further supported potential causal roles for 44 proteins, highlighting their relevance as candidate therapeutic targets. In addition, mediation-prioritized proteins significantly improved the prediction of secondary disease risk beyond conventional clinical factors and other proteomic features. Furthermore, stratification by biomarker-defined disease control and severity revealed additional pathways and mediators, underscoring the importance of disease management in mitigating downstream complications.

Our findings support and extend previous observational studies linking the three primary cardiometabolic diseases--diabetes, hypertension, and dyslipidemia--to a broad range of secondary conditions. Numerous studies have established these disorders as key risk factors for CVDs^3,7,65^ and RDs^30,32^. In addition, our analysis confirmed several well-known disease-protein relationships, including those between diabetes and GDF15 and dyslipidemia and PCSK9.

Our findings further highlight the central role of plasma proteins in linking primary cardiometabolic diseases to downstream complications. Several proteins, including GDF15 and LGALS4, repeatedly mediated associations between a single primary disease and multiple secondary outcomes, whereas others, such as ASGR1, connected all three primary diseases to a common outcome, including heart failure. These broadly distributed mediators may represent shared molecular mechanisms underlying disease progression across organ systems.

Importantly, a subset of proteins appeared to play a more active role in disease development. Mendelian randomization analyses provided evidence supporting potential causal effects for 44 plasma proteins, suggesting that these molecules may contribute directly to pathogenesis rather than simply reflect downstream disease processes. Notable examples included REN, FGF23, and UMOD, all of which demonstrated evidence of causal involvement in renal outcomes. Distinguishing proteins that are reactively elevated during disease progression from those that may causally drive disease development is particularly important, as the latter represent promising candidates for mechanistic investigation and therapeutic intervention.

Prediction analysis demonstrated that the mediation-prioritized proteins provided superior predictive performance for secondary disease risk among individuals with primary diseases, outperforming models based on other plasma proteins or conventional clinical risk factors. For example, improving RTID risk prediction by approximately 12% in individuals with hypertension.

Beyond defining primary disease status based on diagnostic history, we also incorporated continuous clinical biomarkers—HbA1c and blood pressure—to capture disease control and severity, enabling stratification of participants into multiple risk categories. HbA1c-based stratification revealed several novel disease pathways not identified using diagnosis alone, including a previously unreported association between poor glycemic control and increased risk of NRPD. For hypertension, stratification by blood pressure levels yielded disease pathways largely consistent with those identified using diagnostic history, suggesting that quantitative blood pressure measurements recapitulate established risk structures while providing additional resolution of disease severity. These stratifying findings demonstrate the importance of glucose and blood pressure management in reducing secondary disease risk among individuals with cardiometabolic conditions. We did not stratify dyslipidemia severity using biomarkers such as lipoprotein, because dyslipidemia may involve either elevated or reduced lipid levels, depending on disease subtype or treatment status.

In summary, we map the plasma protein mediators that link cardiometabolic disease to its secondary outcomes and define a parsimonious set of candidates with translational potential as biomarkers and therapeutic targets. By resolving the molecular pathways that connect upstream risk factors to downstream complications, this work establishes a framework for clinically accessible markers that can refine risk stratification and guide targeted interventions.

## Limitations of the study

Several limitations of our study should be acknowledged. First, our analysis focused on disease development from three primary cardiometabolic conditions—diabetes, hypertension, and dyslipidemia—to their downstream complications. Although the analytical framework can be readily extended to other disease pathways, our findings are limited to these cardiometabolic contexts. Second, although pathway enrichment analyses and two-sample Mendelian randomization provided biological context and supported potential causal roles for a subset of mediator proteins, these findings should primarily be interpreted as statistically prioritized candidates rather than definitive mechanistic discoveries. Our future work will focus on integrating experimental validation and computational approaches, such as network-based causal hierarchy analysis, a comprehensive colocalization analyses, and cross-organ interaction modeling, to further clarify the biological mechanisms underlying these mediation pathways. Third, medication use among individuals with prevalent primary diseases may have influenced both protein levels and disease risk. While we conducted biomarker based stratified analysis to address this issue, residual confounding related to treatment exposure cannot be excluded. Lastly, the generalizability of our findings may be limited, as the majority of participants in the UK Biobank cohort were white individuals of European ancestry. Further validation in more diverse populations will be necessary to determine whether these mediation pathways are consistent across different genetic and environmental backgrounds.

## Methods

### Experimental model and study participant details

Plasma proteomic measurements were obtained from the UK Biobank Pharma Proteomics Project (UKB-PPP)^21^, which used the Olink Explore 3072 proximity extension assay (PEA) platform, an antibody-based technique. We included 53,030 participants in our study, whose blood samples were collected between March 2006 and September 2010 and measured between April 2021 and February 2022, yielding 2,941 protein analytes corresponding to 2,923 unique proteins. The time point of plasma protein data collection was defined as instance 0. A subset of participants underwent two additional rounds of plasma collection from 2015 to 2019 and again in 2021. However, these profiles covered a smaller panel of proteins (1,173), with only 506 proteins overlapping with those collected at instance 0. Due to this limited overlap, these later profiles were not included in the primary analyses. In addition, GLIPR1 was excluded due to insufficient data availability, resulting in 2,922 plasma proteins included in the final analyses. As one of the primary objectives of this study was to evaluate the predictive performance of selected plasma proteins for secondary disease risk, we randomly assigned 80% of participants to a training set for mediation analysis and predictive model development, and assigned rest of participants to test set.

Participant age was defined as age at the UK Biobank assessment center. Sex was obtained from UK Biobank records, which were acquired from central registry information at recruitment and could be updated by participants. Ethnic background was based on self-reported UK Biobank assessment data. Socioeconomic status was represented by the Townsend deprivation index at recruitment.

### Primary diseases and secondary diseases

The primary diseases analyzed in this study included diabetes, hypertension, and dyslipidemia, which are well-known risk factors for multiple subsequent diseases. In this study, we examined secondary diseases across four major disease categories, including cardiovascular, pulmonary, renal, and neurological disease. After excluding disease subtypes with very low prevalence (< 0.1%), a total of 18 secondary diseases were included in the analysis. Disease categorization followed the original UK Biobank disease classification framework and was further reviewed by clinical expertise. Detailed categorization, corresponding ICD-10 codes, and disease prevalence are provided in Table S3.

Primary disease status was defined as a binary variable, indicating whether participants were diagnosed with a primary disease within a window from two years before to one year after the plasma protein collection (instance 0). Individuals diagnosed more than two years before instance 0 were excluded to ensure that primary disease status closely reflected the time point of plasma protein collection. The number of cases for each primary disease ranged from 1,053 (2.0%) to 3,012 (5.7%).

Participants diagnosed with secondary diseases prior to instance 0 or before the onset of the primary disease were excluded, as the analysis focused on the future risk of secondary disease development. Follow-up time and outcome information were obtained from the UK Biobank longitudinal health records. For each outcome, follow-up began at instance 0 and continued until the earliest occurrence of the outcome event or the end of follow-up (September 1, 2023).

### Covariates

The covariates considered in this study included participants’ age, sex, race, body mass index (BMI), smoking status, alcohol consumption status, diastolic blood pressure (DBP), systolic blood pressure (SBP), glycated hemoglobin (HbA1c) and low-density lipoprotein (LDL). These variables are well-established risk factors and potential confounders for cardiometabolic and chronic diseases^70–72^, including cardiovascular, renal, neurological, and pulmonary diseases. Information on smoking status and alcohol consumption was collected through questionnaires administered at baseline (instance 0). Smoking status was categorized into three levels (never, past, and current), and alcohol consumption was categorized into five levels (never, monthly or less, 2–4 times per month, 2–3 times per week, and ≥4 times per week). Measurements of SBP, DBP, and HbA1c were also obtained at baseline. To avoid over-adjustment, covariates that directly overlapped with the definitions of primary diseases were excluded from certain models. For example, SBP and DBP were excluded when hypertension was the primary disease, whereas HbA1c and LDL were retained to account for potential confounding by diabetes and dyslipidemia.

### Stratification by blood pressure and glycated hemoglobin

In addition to defining primary disease status based on diagnostic history, we further stratified participants according to SBP, DBP, and HbA1c levels. Unlike historical disease data, blood pressure and glycated hemoglobin measurements were obtained on the same day as the plasma protein data collection (instance 0). However, these physiological markers may be more sensitive to short-term environmental or individual factors, such as medication use and recent physical activity. Therefore, blood pressure and glycated hemoglobin levels were used as complementary indicators of disease severity and control alongside diagnostic history.

According to the 2023 European Society of Hypertension (ESH) guidelines^43^, participants were classified into four blood pressure categories: normal (< 140/90 mmHg), hypertension grade 1 (140–159/90–99 mmHg), hypertension grade 2 (160–179/100–109 mmHg), and hypertension grade 3 (≥ 180/110 mmHg). Similarly, participants were classified based on HbA1c levels^38^ into normal (HbA1c < 5.7%), prediabetes (5.7% ≤ HbA1c < 6.4%), and severe-diabetes (HbA1c ≥ 6.4%).

Using these categorizations, we examined whether participants with elevated blood pressure (grades 1 to 3) or impaired glycemic control (prediabetes and severe-diabetes) exhibited an increased likelihood of developing secondary diseases compared to those with normal levels. Mediation analysis was then performed to assess the role of plasma proteins in mediating associations between disease severity or control and secondary disease risk.

### Mediation analysis

Under the mediation analysis framework^21^, we defined the onset of primary disease as the exposure variable, baseline expression levels of 2922 plasma proteins as potential mediators, and the occurrence and timing of secondary diseases as outcome variables. All analyses were conducted exclusively within the training set. We first evaluated the association between primary disease status and secondary disease risk using a Cox proportional hazards model^73^,

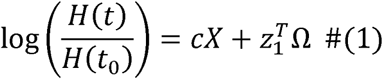

where *H(t)* denotes the hazard function at time t, *t*_0_ is the time at the instance 0, *t*_0_ is the log relative hazard (defined as the total effect), *X* is the binary primary disease status (diagnosed vs. not diagnosed), Ω is the vector of covariates, and, *z*_1_ denotes the corresponding coefficients.

Covariates were described in the Covariate section. Secondary diseases showing statistically significant associations with primary diseases (Bonferroni adjusted *p* < 0.05) were advanced to subsequent analyses.

Next, we assessed the associations between plasma protein levels and primary disease status using linear regression models with the same covariate set,

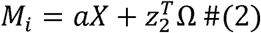

where, *M_i_*, is the expression level of protein *i a* represents the protein expression change associated with primary disease status, and *z*_2_ represents the coefficients for covariates. Proteins were considered significantly associated with the primary disease if the Bonferroni adjusted *p*-values were < 0.05 and were subsequently advanced to further analyses.

Finally, we examined the potential mediating role of plasma proteins using a Cox model incorporating both exposure and mediator variables,

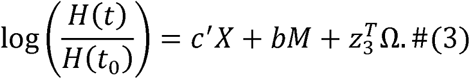

where *c*^J^ represents the direct effect of the primary disease, *b* represents the effect of one plasma protein, and *z*_3_ epresents covariate effects. The indirect effect of each protein was estimated using the product-of-coefficients method^21^ (*a b*) and its statistical significance was evaluated via 1,000 bootstrap resamples^74^. Resulting *p* -values were adjusted suing the Bonferroni procedure. This analytical pipeline was applied for each disease pathway.

Plasma proteins were defined as significant mediators if they met all the following criteria: (i) a statistically significant indirect effect after multiple testing correction, (ii) an absolute value of the indirect effect ≥ 0.01, and (iii) a proportional indirect effect (PIE), define as the ratio of the indirect to the total effect, of at least 10%. We further defined key protein mediators as those that: (i) consistently mediated pathways from a single primary disease to multiple secondary diseases; (ii) exhibited strong mediation effects within individual disease pathways; or (iii) mediated pathways that linked all three primary diseases to a common secondary disease. We further evaluated the association between the significantly mediated plasma proteins and the risk of secondary diseases using Cox proportional hazards models adjusted for the same set of covariates. Resulting *p*-values were adjusted suing the Bonferroni procedure.

### Mendelian randomization

To evaluate the potential causal effects of identified plasma mediators on their corresponding secondary diseases, we performed two-sample Mendelian Randomization (MR)^75^. We used *cis*-pQTLs as instrument variable (within 1 Mb of the transcription start site) from the UKB-PPP at genome-wide significance (*p* □ < □5□×□□10)^23^. Independent instruments were selected through linkage disequilibrium (LD) clumping (r² < 0.01) using the European reference panel, discarding instruments with F-statistics<10 to mitigate weak instrument bias. Genetic associations of the selected variants with secondary diseases were extracted from publicly available genome-wide association study (GWAS) summary statistics from the IEU OpenGWAS database^55,66^, restricted to individuals of European ancestry. Harmonization procedure was performed to align effect alleles across exposure (pQTLs) and disease outcome GWAS datasets, and palindromic variants with ambiguous strand orientation were excluded.

Twelve MR methods, including inverse-variance weighted (IVW) method (fixed effect, multiplicative random effects with/without corrected standard error under dispersion), MR-Egger regression, Simple median, Weighted median, Penalised weighted median, Simple mode, Simple mode (NOME), Weighted mode, Weighted mode (NOME), were applied^57^. For single-variant instruments, the Wald ratio was used. Heterogeneity across instruments was assessed using Cochran’s Q statistic and the I² metric, and horizontal pleiotropy was evaluated using the MR–Egger intercept. An association was considered to show a potential causal relationship if the Bonferroni adjusted *p-*value was <0.05 in at least one MR method. All analyses were conducted using the TwoSampleMR package in R^55^.

### Predicting the future risk of secondary diseases

To evaluate the predictive power of identified plasma protein mediators, we applied a LASSO-penalized Cox survival model^62,68^ using the top ten proteins with the largest indirect effects as predictors. For each primary–secondary disease pathway, models were trained using subsets of participants with the corresponding primary disease in the training set, and predictive performance was assessed in the independent test subset using the concordance index (C-index)^64^ to quantify out-of-sample discrimination ability among participants with the same primary disease. Prediction models hyperparameters were selected by 10-folds CV within the training subset.

The prediction performance of this mediation-informed model was compared with four alternative approaches: (1) a LASSO survival model including only basic demographic variables (age, sex, BMI, and race); (2) a LASSO survival model incorporating conventional clinical and lifestyle risk factors, including age, sex, BMI, blood pressure, HbA1c level, smoking and drinking statues; (3) a LASSO survival model using ten randomly selected plasma proteins, with the random selection repeated 100 times to obtain the average C-index; (4) an XGBoost survival model^69,76^ using all 2,922 plasma proteins as predictors. Due to the substantial proportion of missing values when including all plasma proteins (10.3% missingness), LASSO-based models were not suitable. Therefore, we employed an XGBoost-based model, which is more robust to missing data. For comparability, all models used identical training–test splits defined by a fixed random seed.

### Sensitivity analysis

To evaluate the robustness of proteins identified through mediation analysis, we performed a sensitivity analysis with additional adjustment for 10 clinical and biochemical covariates. These included markers of hepatic function (alanine aminotransferase and aspartate aminotransferase), hematologic parameters (mean corpuscular volume, hemoglobin concentration, and platelet count), renal function indicators (cystatin C and creatinine), metabolic markers (total cholesterol and albumin), as well as socioeconomic status measured by the Townsend deprivation index (TDI). We then compared the top five proteins (ranked by indirect effect) identified for each disease pathway between the primary covariate model and the extended model including these additional covariates, in terms of Jaccard index^37^.

### Biological function enrichment analysis

Biological processes and pathways were assessed using R^67^ package clusterProfiler (v4.16.0)^53^, drawing on Gene Ontology Biological Process and KEGG. Plasma proteins were mapped to human gene identifiers using org.Hs.eg.db (v3.21.0) annotation database, retaining unique mapped genes and excluding unmapped entries^77^. Enrichment analysis was conducted with the function enrichGO(·) and enrichKEGG(·). The background gene set was defined as all 2,922 proteins quantified in the proteomic platform, to control for platform coverage. Biological were considered significantly enriched if the Benjamini-Hochberg (BH) adjusted *p*-values^78^ were < 0.1. To reduce redundancy among enriched GO terms, semantic similarity analysis was performed using the simplify(·) function in the clusterProfiler package. GO terms with a pairwise semantic similarity greater than 0.7 were considered redundant, and the term with the lowest BH adjusted *p*-value was retained as the representative term.

## Data and code availability

All UK Biobank data (plasma proteome profile and longitudinal clinical data) are accessible via application to the UK Biobank. PQTL summary statistics from the UKB-PPP can be accessed at https://metabolomips.org/ukbbpgwas. All analyses and visualization were implemented in R (v4.4.0). Scripts are available on GitHub: https://github.com/mengwang-lab/Plasma-protein-mediation-analysis. Any additional information required to reanalyze the data reported in this work paper is available from the Lead Contact upon request.

## Data Availability

All UK Biobank data (electronic health record, plasma proteome profile) are accessible via application to the UK Biobank. PQTL summary statistics from the UKB-PPP can be accessed at https://metabolomips.org/ukbbpgwas.

## Acknowledgements

M.W., and H.Q., was supported by M.W.’s startup fund from the Institute for Heart and Brain Health. This work was supported by grants the NIH R35 HL155318 to A.R. and the American Heart Association (AHA) grants 23MERIT1038415 to A.R. and SFRN (URLs: https://doi.org/10.58275/ AHA.24SFRNPCN1284382.pc.gr.194135 and https://doi.org/10.58275/ AHA.24SFRNCCN1276092.pc.gr.194131) to A.R. and M.W.

## Author contributions

M.W. conceived the study, supervised the research, and revised the manuscript. A.R. supervised the research and revised the manuscript. H.Q. performed the analyses and wrote the manuscript. C.W. supported the pathway and biological process analyses and revised the manuscript. B.L. assisted with figure preparation and revised the manuscript. All authors had access to the data used in this study; H.Q., C.W., and M.W. had unrestricted access to all data and verified the underlying data. All authors agreed to submit the manuscript, read and approved the final draft, and take full responsibility for its content.

