## Supplementary Materials for "Plasma Proteomics Linking Primary and Secondary diseases: Insights into Molecular Mediation from UK Biobank Data" (from another version of this preprint: version 2, 2026-04-19)

**Diabetes Control–Based Mediation Results**

To assess whether participants with different levels of diabetes exhibit varying risks of secondary diseases and corresponding mediated proteins, glycated hemoglobin (HbA1c) was further examined in relation to differential risks of secondary diseases and distinct protein-mediated pathways. Participants were stratified according to HbA1c levels^1^: normal (HbA1c < 5.7%), prediabetes (5.7% ≤ HbA1c < 6.4%), and severe diabetes (HbA1c ≥ 6.4%).

We observed a substantial number of mediating proteins associated with prediabetes (Supplementary Table 10 and Supplementary Table 11). Specifically, 394 proteins mediated the relationship between prediabetes and 10 secondary diseases, exceeding the number identified for severe diabetes (352 proteins linked to 12 secondary diseases) and for diabetes defined by diagnosis history (341 proteins), with 199 proteins shared across these classifications (Supplementary Fig 4a-b). When comparing the magnitude of indirect effects for overlapping proteins linked to the same secondary diseases, severe diabetes consistently exhibited stronger mediation effects, followed by diabetes diagnosis history (Supplementary Fig. 4c).

As we found the importance of blood glucose management in reducing secondary disease incidence, we further evaluated an optimal HbA1c control level to prevent secondary disease. We used a generalized additive model (GAM) to assess the association between HbA1c levels and the risk of multiple secondary diseases^2^. For AD, we observed a sharp increase in risk when HbA1c levels exceeded approximately 5.7-5.8%, with a steeper rise beyond 7.9% (Supplementary Fig. 4e), which aligns with the clinical practice and enhances the importance of maintaining normal blood glucose (HbA1c < 5.7%), the secondary disease risk dramatically increases even in the moderate blood glucose range. In contrast, risk remained relatively low at HbA1c levels below 5.7%. Additionally, uncertainty, reflected by wider confidence intervals, increased markedly at extremely high HbA1c values, underscoring the importance of maintaining glycemic control within a normal range to reduce the risk of adverse outcomes. These results suggest the presence of two critical HbA1c “acceleration points”, around 5.7% and 7.9%, beyond which the risk of AD increases substantially.

A distinct pattern was observed for valve disorders, where the lowest risk occurred near an HbA1c level of 5.2%, followed by a pronounced increase in risk beyond approximately 5.8% (Supplementary Fig. 4f). Interestingly, a modest elevation in risk was also noted at very low HbA1c levels; however, this may be attributed to statistical variability at the extremes of the HbA1c distribution, suggesting that both hyperglycemia and hypoglycemia warrant further investigation. For most secondary diseases, we observed a largely linear relationship between disease risk and HbA1c levels within the intermediate range, reinforcing the clinical relevance of HbA1c as a continuous biomarker for disease risk stratification.

**Hypertension Control–Based Mediation Results**

To assess whether participants at different levels of blood pressure have different risks for secondary diseases, as well as distinct patterns of protein mediators along these disease pathways, we further stratified participants by their systolic and diastolic blood pressure (SBP and DBP) into four categories: normal (< 140/90 mmHg), hypertension grade 1 (140–159/90–99 mmHg), hypertension grade 2 (160–179/100–109 mmHg), and hypertension grade 3 (≥ 180/110 mmHg). This stratification approach was based on the 2023 European Society of Hypertension (ESH) guidelines^3^ and is described in detail in the Methods section.

A total of 197 plasma proteins were identified as mediators in the relationship between at least one grade of hypertension severity and a secondary disease (Supplementary Table 8 and Supplementary Table 9). Participants with grade 3 hypertension were more likely to exhibit protein-mediated increases in secondary disease risk compared to those with lower hypertension levels, as evidenced by both a broader range of mediating proteins (162 unique proteins, Supplementary Fig. 6a) and stronger indirect effects through overlapping plasma proteins linked to the same secondary diseases (Supplementary Fig. 6b). Many of the key mediators linked to grade 3 hypertension overlapped with those identified when hypertension status was defined based on diagnostic history (Supplementary Table 9). For example, HAVCR1 was associated with the largest number of secondary diseases (n = 6) and also exhibited strong mediation effects, while MMP7 demonstrated significant indirect effects across five disease mediation pathways under both classification strategies. Additionally, NT-proBNP exhibited particularly strong mediation effects for several cardiovascular outcomes, including HF (HR = 1.16, PIE = 38%) .

Nevertheless, we also identified proteins uniquely associated with grade 3 hypertension that were not detected when using diagnostic history alone. For example, CRIP2, a protein highly expressed during cardiovascular development, emerged as a significant mediator in the grade 3 hypertension analysis but not under the diagnosis-based definition. Furthermore, comparative analyses of overlapping proteins linked to the same secondary diseases revealed that proteins identified through grade 3 hypertension consistently exhibited stronger indirect effects than those identified via hypertension diagnosis (Supplementary Fig 6c). Notably, several of these newly identified mediators also showed evidence of potential causal effects on secondary diseases, as supported by Mendelian randomization analysis.

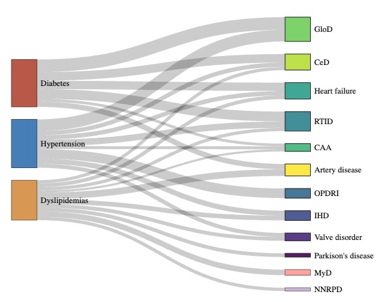

Supplementary Fig. 1. Disease development pathways between three primary diseases and secondary diseases in 4 clinical domains. The width of the line represents the hazard ratio.

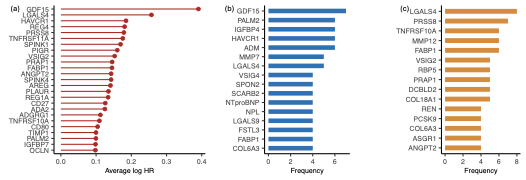

Supplementary Fig. 2. Plasma proteins exhibiting mediation effects across multiple disease pathways. (a) Average indirect effects (log hazard ratios) of 25 plasma proteins that mediated all six diabetes-related disease pathways. (b) Number of hypertension-related disease pathways in which each plasma protein exhibited a significant mediation effect. (c) Number of dyslipidemia-related disease pathways in which each plasma protein exhibited a significant mediation effect.

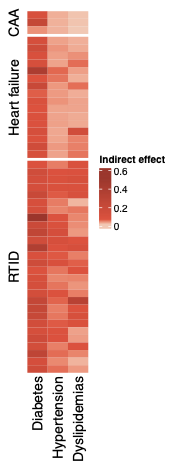

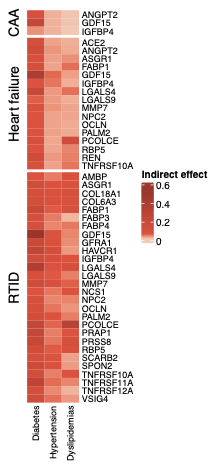

Supplementary Fig. 3. The mediation effect of proteins that consistently link all three primary diseases to one secondary disease.

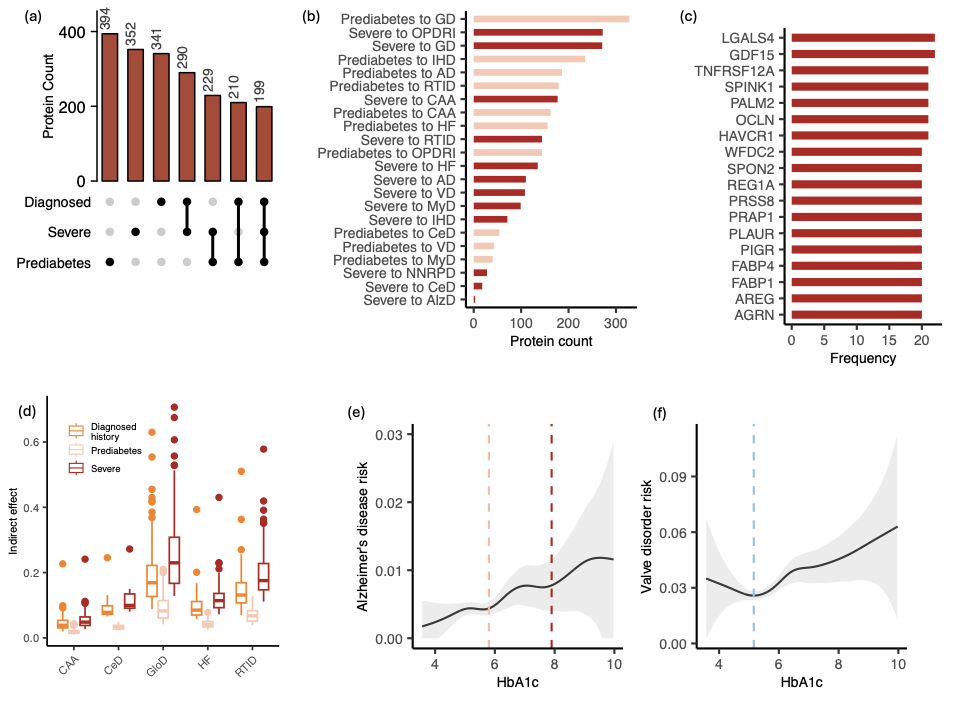

Supplementary Fig 4. Mediation analysis results stratified by diabetes control levels based on HbA1c. (a) Number of plasma proteins mediating each diabetes-to-secondary disease pathway under different diabetes grades. (b) Number of plasma proteins mediating each disease path across different diabetes severity levels. (c) Frequently observed protein mediators that consistently link varying levels of diabetes severity to diverse secondary diseases. (d) Comparison of the mediation effects of proteins across different diabetes grades. (e, f) Nonlinear associations between HbA1c levels and the risk of Alzheimer’s disease (e) and valve disorders (f), estimated using generalized additive models. For Alzheimer’s disease, the vertical lines indicate two inflection points beyond which disease risk increases rapidly. For valve disorders, the vertical line marks the HbA1c level associated with the lowest estimated risk.

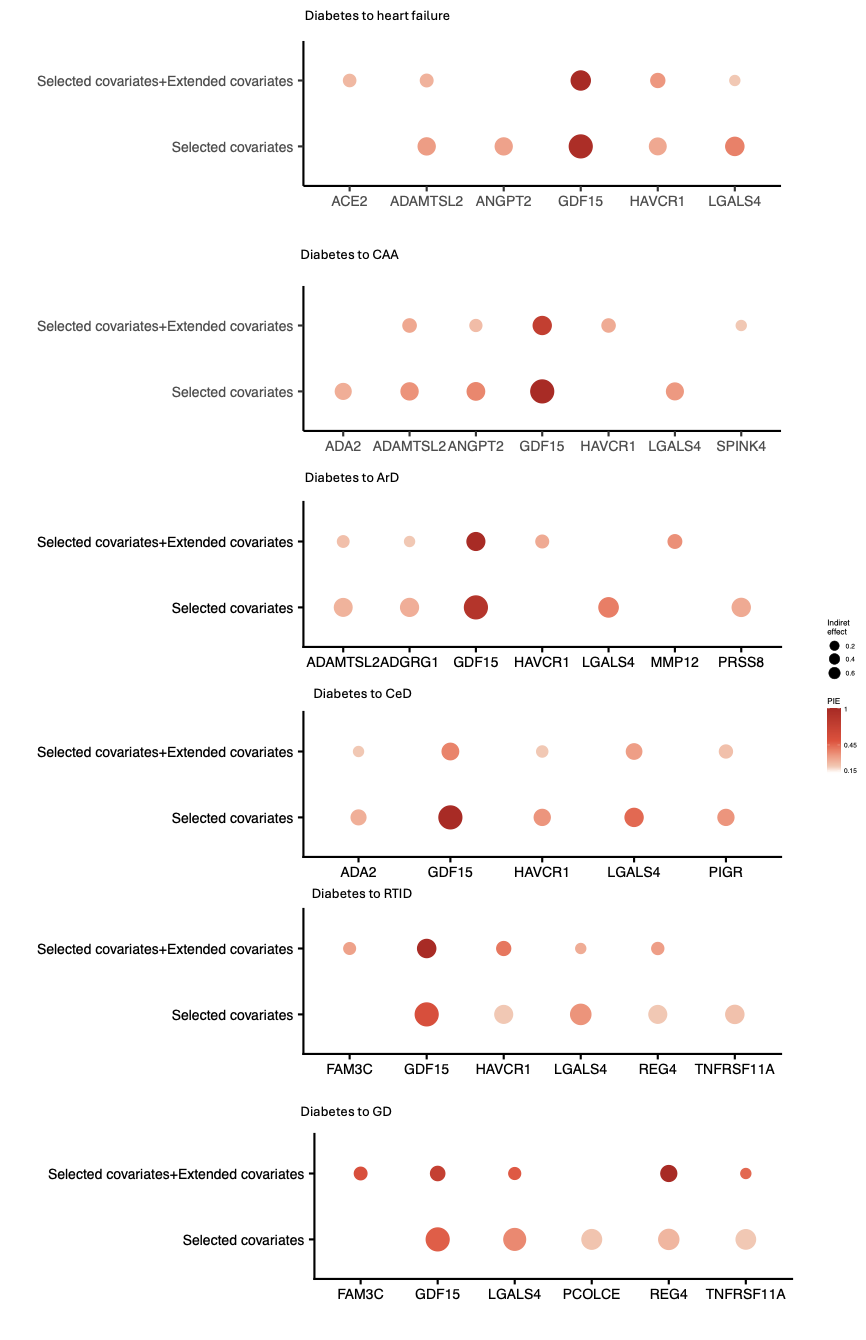

Supplementary Fig. 5. Comparison of the top five mediating proteins identified using the base covariate model versus the extended covariate model.

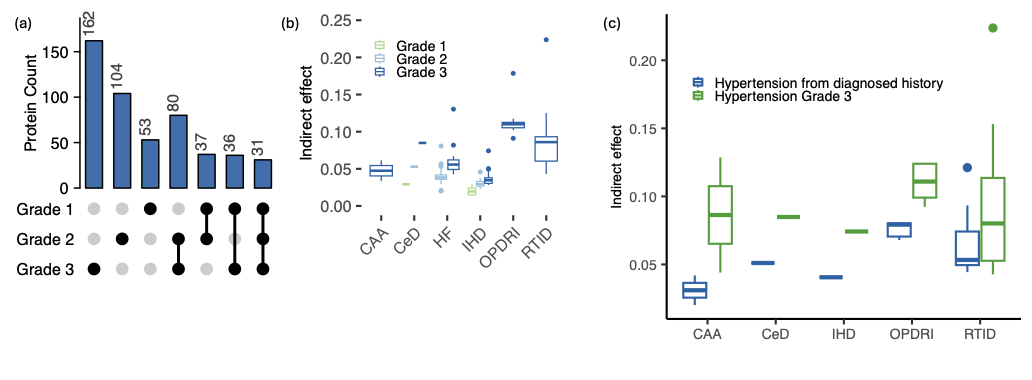

Supplementary Fig 6. Mediation analysis results stratified by hypertension control levels based on blood pressure. (a) Number of plasma proteins mediating each grade-specific hypertension–disease path. (b) Comparison of the mediation effects of proteins across different hypertension grades. (c) Comparison of mediation effects for the proteins linking to the same secondary disease, evaluated separately for grade 3 hypertension and binary hypertension diagnosis.

1.	ElSayed, N. A. *et al.* 2. Diagnosis and Classification of Diabetes: Standards of Care in Diabetes—2025. *Diabetes Care* **48**, S27–S49 (2025).

2.	Wood, S. N. *Generalized Additive Models: An Introduction with R, Second Edition*. *Generalized Additive Models: An Introduction with R, Second Edition* (2017). doi:10.1201/9781315370279.

3.	Mancia, G. *et al.* 2023 ESH Guidelines for the management of arterial hypertension The Task Force for the management of arterial hypertension of the European Society of Hypertension. *J. Hypertens.* **41**, 1874–2071 (2023).
